# Improvement in Chronic Kidney Disease (CKD) diagnosis and management after automatic loading of CKD in the electronic health record problem list

**DOI:** 10.64898/2026.11.12.26367913

**Authors:** Waleed Zafar, Chen Dun, Yirui Hu, Lauren Brubaker, Celia Gray, Cory Siegrist, H. Les Kirchner, Jamie Green, Jung-Im Shin, Alexander R. Chang

**Author notes:** **Corresponding Author:** Alexander Chang, MD, MS, 100 N Academy Ave, Danville, PA 17822.

## Abstract

**Background:** Chronic kidney disease (CKD) is often underrecognized and undiagnosed. We evaluate the impact of an algorithm-based automatic loading process for CKD in the electronic health record (EHR) problem list at a large health system.

**Methods:** Study populations included patients receiving primary or nephrology care at Geisinger (intervention group) and 4 large health systems in the Optum Labs Data Warehouse (OLDW), a longitudinal, real-world data asset with de-identified administrative claims and electronic health record (EHR) data,(comparator group) from 2008-2023. Since 2013, an automated process has been regularly implemented at Geisinger, adding CKD to problem lists of patients meeting laboratory criteria for CKD (outpatient eGFR <60 ml/min/1.73m^2^ on 2 lab values for ≥ 3 months without intervening normal values). We conducted difference-in-differences (DiD) and difference-in-difference-in-differences (DiDiD) analyses comparing CKD diagnosis rates, CKD-related lab tests, use of renin-angiotensin-aldosterone system inhibitors (RAASi) and statins for those >50 years within 1 year of incident CKD (index date) at Geisinger vs. OLDW health systems before (2008-2012) and after (2013-2023) CKD auto-load implementation. Analyses relied on summary-level data and did not permit adjustment for individual-level characteristics.

**Results:** Among patients with incident, undiagnosed CKD (n=21,995 in Geisinger; n=208,402 in OLDW), CKD diagnosis within 1 year of index date increased from 28.4% in 2008-2012 to 65.7% in 2013-2023 at Geisinger whereas CKD diagnosis decreased from 22.6% to 20.7% at OLDW (DiD estimate 39.1%; 95% CI: 38.1%, 40.1%; p<0.001). Among CKD-related labs, DID estimates favored Geisinger for follow-up eGFR, parathyroid hormone, and phosphorus testing (all p values <0.001). CKD auto-load was associated with increased use of RAASi and statins (p values <0.05).

**Conclusions:** An algorithm-based problem list auto-load process was associated with higher CKD diagnosis, some CKD-related labs, and use of recommended medications. This scalable strategy should be validated in other health systems.

## Introduction

Chronic kidney disease (CKD) affects 1 in 7 U.S. adults^1^ and significantly increases risks of cardiovascular disease and death,^2^ yet as few as 1 in 5 individuals with CKD are aware of their diagnosis.^3^ The documentation of CKD in the electronic health record (EHR) also remains suboptimal thereby reducing its recognition and management in primary care as well as making it a patient safety issue.^4^ Prior studies report that fewer than 1 in 5 individuals who meet laboratory diagnosis criteria have CKD documented as a problem^5 6^ and on average, it takes several years before CKD is diagnosed.^7^

The patient’s “problem list” within the EHR has been suggested as an effective way to improve CKD awareness and its management at the primary care.^8 9^ The EHR problem list can help clinicians quickly organize clinical information, which can then help providers focus on medical conditions during clinical visits, order appropriate workup and treatment; the problem list is often used to trigger clinical decision support (CDS) tools to improve disease management.^6, 9-11^ The problem list is not encounter-specific but rather remains constant across different types of clinical encounters, can feed into the assessment and plan section of clinical notes, and can be visible to patients through electronic patient portals.^11^

In this study, we evaluate the impact of a novel strategy to improve CKD recognition through the use of algorithm-based automatic loading of CKD in the EHR problem list among primary care patients who met laboratory diagnosis of incident CKD at a large regional health system.

## Methods

### Setting

Geisinger is a large, mostly rural health system in central and northeast Pennsylvania providing care to approximately 1.2 million people. A CKD auto-loading process was developed at Geisinger and has been running on a regular basis (approximately monthly) to identify patients with undiagnosed CKD since 2013. Patients were identified based on two outpatient eGFR values <60 ml/min/1.73m^2^, measured at least 3 months apart with no normal values in between. The process then automatically adds CKD diagnosis to the patients’ problem list. **Figure 1** shows how the Geisinger Epic^®^ EHR displays CKD diagnosis when it has been autoloaded.

**Figure 1:**
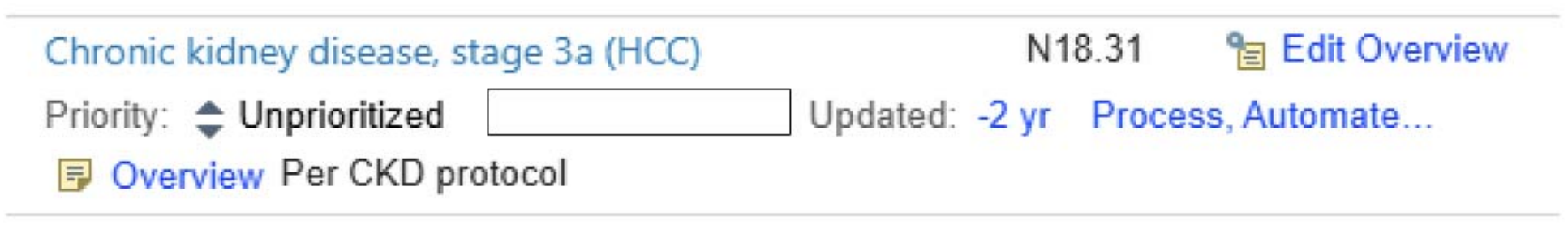
Example of Electronic health record documentation of “autoloaded” CKD diagnosis in the problem list

We conducted a retrospective analysis of Geisinger patients receiving primary care or nephrology and evaluated the presence of CKD as a diagnosis during outpatient visit in the 1 year after fulfilling laboratory diagnostic criteria for CKD (index date). We compared the results with data from 4 health systems similar in size to Geisinger in the Optum Labs Data Warehouse (OLDW), a longitudinal, real-world data asset with deidentified administrative claims and EHR data.^12^ The rationale for selecting the similar or larger-sized health systems was to enhance comparability and reduce influence of sparse data from smaller health systems. This study was approved by the Geisinger Institutional Review Board.

### Study population

The eligible population comprised all adults ≥18 years actively receiving primary care or nephrology at Geisinger or OLDW (defined as having received a primary care provider [PCP] or nephrology appointment in the health system within 2 years before the index date) who met laboratory diagnostic criteria for incident CKD (eGFR <60 ml/min/1.73m^2^ [CKD-EPI 2021 equation]^13^ on 2 labs separated by at least 3 months but no more than 2 years, with no normal eGFR value ≥60 between the 2 qualifying labs). Index date was defined as the date of last outpatient eGFR value qualifying for incident CKD. Patients were excluded if they had received CKD diagnosis code (ICD-9 or ICD-10 codes for chronic kidney disease, diabetic nephropathy, unspecified renal failure, hypertensive kidney disease) in any encounter or on problem list prior to the index date, were under hospice care, were receiving palliative care or had existing diagnosis of end-stage kidney disease (ESKD).

#### Outcomes

The primary outcome was presence of a CKD ICD-9 or 10 code as a diagnosis during outpatient visit in the 12 months after the index date. We also examined secondary outcomes including completion of CKD-related labs (i.e. creatinine, eGFR, albumin/creatinine ratio (ACR), protein/creatinine ratio (PCR), intact parathyroid hormone, phosphorus, 25-hydroxyvitamin D, hemoglobin, lipid panel) within 12 months after the index date. We also evaluated change in CKD-related guideline directed medical therapy (GDMT) use within 12 months after the index date (prescription of angiotensin-converting enzyme inhibitors [ACEi] or angiotensin-receptor blockers [ARB], sodium-glucose cotransporter 2 inhibitors [SGLT2i], statins for those above the age of 50 years).^14^ We did not include glucagon-like peptide 1 receptor agonists (GLP-1 RA) or nonsteroidal mineralocorticoid receptor antagonists (nsMRA) in these analyses given the later appearance of these in guidelines and very low uptake of nsMRA overall, consistent with previously published work.^15^

#### Statistical analyses

Descriptive statistics were used to summarize baseline characteristics at Geisinger and the 4 OLDW health systems. Continuous variables are reported as mean and standard deviation (SDs), and categorical variables as counts and percentages. Temporal trends in CKD diagnosis code documentation alongside CKD problem list auto-loading at Geisinger were evaluated descriptively. The association between yearly CKD auto-loading and CKD diagnosis code documentation was evaluated using Pearson correlation coefficients and linear regression analyses. Time- to-event analyses using Kaplan-Meier curves were done to understand differences in time to CKD diagnosis between CKD stages 3a, 3b, and 4 for both cohorts.

A difference-in-differences (DiD) design was used to estimate the effect of CKD auto-loading while accounting for secular trends in CKD diagnosis, using the 4 OLDW health systems as a non-intervention comparator. Because detailed implementation records were unavailable, we retrospectively characterized CKD problem list auto-loading by identifying CKD problem list entries that were accompanied by a note indicating that it was automatically generated (i.e. “Per CKD protocol”) versus those that were added manually (**Figures 1,2**). This process was evident as early as 2008, when 3% of adults meeting lab diagnostic criteria for incident CKD had an auto-loaded CKD problem list diagnosis. Use of the process increased but appeared inconsistently implemented from 2009 to 2012 (23% to 41% of incident CKD patients had auto-loaded CKD problem list diagnoses). Auto-loading increased substantially to 58% in 2013 and then remained high (range 68% to 87%) each year through 2023, consistent with broad sustained implementation. Comparable problem list data were unavailable within OLDW. Therefore, we defined the time period 2008-2012 as the pre-implementation period and 2013-2023 as the post-implementation period. Then, we compared changes in CKD diagnosis documentation at Geisinger at outpatient visits with contemporaneous changes across the 4 OLDW health systems.

**Figure 2.**
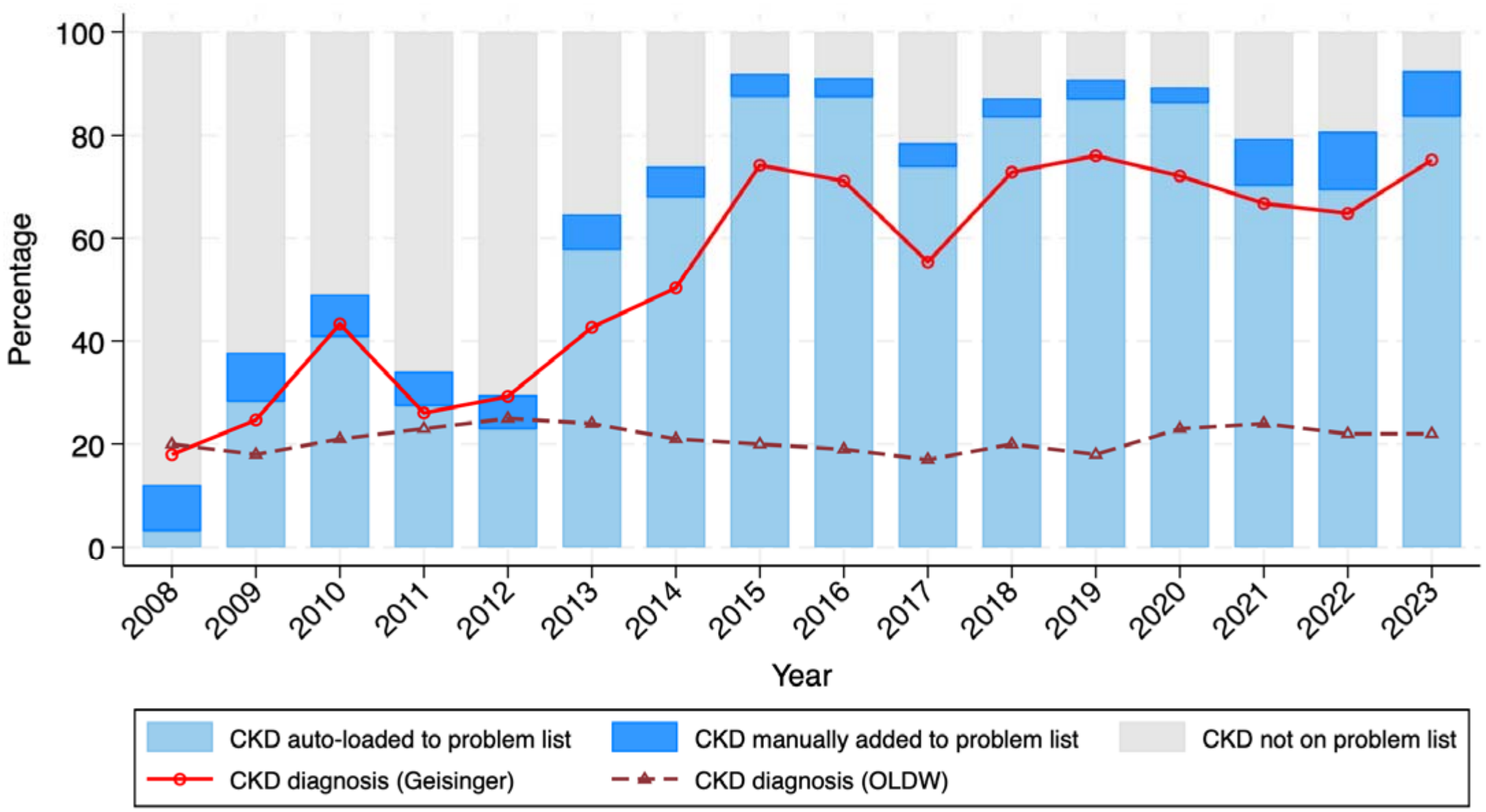
Trends in CKD Problem List at Geisinger and CKD Diagnosis at Geisinger vs. 4 OLDW Health Systems As shown in the figure, CKD problem list auto-loading was occurring in a minority of patients prior to 2013 and then was implemented much more regularly from 2013 onwards at Geisinger. Trendlines for CKD diagnosis at outpatient encounters within 1 year of index date of incident CKD (based on lab values) are shown for Geisinger and 4 comparator OLDW health systems.

To evaluate changes in CKD-related lab monitoring following the index date, we performed DiD analyses comparing changes in proportion of incident CKD patients receiving lab testing within 12 months of index date during the post vs. pre-implementation periods at Geisinger relative to the corresponding changes in the 4 OLDW health systems. To evaluate the association of CKD auto-loading with GDMT use, we controlled for pre-CKD diagnosis baseline medication use, by performing difference-in-difference-in-differences (DiDiD) analyses based on aggregated event counts and denominators. Medication use was compared during the 12 months before and after the index date of incident lab-defined CKD. For ACEi/ARB use and statin use (among adults aged >50 years), we compared the pre vs. post-implementation periods at Geisinger relative to corresponding periods across the 4 OLDW health systems. We fit grouped binomial logistic regression models with terms for health system (Geisinger vs OLDW), implementation period (2013-2023 vs. 2008-2012), assessment window (post-index vs. pre-index), and the three-way health system x implementation period x assessment window interaction. The exponentiated coefficients for the three-way interaction were reported as the DiDiD odds ratio (OR) with Wald-based 95% confidence intervals and p-values. This estimate represents whether change in medication use after vs. before incident lab-based CKD differed between the auto-load time period and the pre-implementation time period at Geisinger beyond temporal changes observed in OLDW.

SGLT2i was evaluated separately as an exploratory analysis since these medications were not available during the original 2008-2012 pre-implementation time period. The first SGLT2i was FDA-approved in 2013, and it was not until 2020 that SGLT2is were explicitly recommended for a CKD indication when the American Diabetes Association (ADA) and the Kidney Disease Improving Global Outcomes (KDIGO) recommended SGLT2i for diabetic kidney disease.^16,17^ Thus, we defined 2014-2020 “pre-guideline era” and 2021-2023 “post-guideline era”. The model used the same specifications as the ACEi/ARB and statin analyses but used these time periods instead. Therefore, the three-way interaction assessed whether the pre- to-post incident lab-based CKD change in SGLT2i use differed between 2021-2023 and 2014-2020 at Geisinger relative to corresponding change in OLDW.

Analyses relied on summary-level data and did not permit adjustment for individual-level baseline characteristics. Two-sided p<0.05 was considered statistically significant. Analyses were performed in R (version 4.4.3) and Stata/MP 19.0.

## Results

A total of 21,995 individuals at Geisinger and 208,402 individuals at the 4 health systems within OLDW were included (**Table 1**). Mean age was 71 (SD 10) years at Geisinger and 69 (SD 11) years in the OLDW health systems. There was a higher proportion of females than males in both the populations (61% and 59% for Geisinger and OLDW). Overall, the population at Geisinger had higher proportions of Whites and commercial insurance coverage and a lower proportion receiving Medicaid. The Geisinger population had higher proportions of individuals diagnosed with diabetes, myocardial infarction, stroke, and heart failure but lower proportions with coronary artery disease and peripheral artery disease than the 4 OLDW health systems (some of these differences likely result from coding differences across health systems).

**Table 1:** Baseline characteristics at Geisinger and in the 4 health systems of similar size or greater than Geisinger within the Optum Labs Data Warehouse data.

|  | **Geisinger**<br>**N = 21,995** | **OLDW**<br>**N = 208,402** |
|---|---|---|
| **Age; Mean (SD)** | 71 (10) | 69 (11) |
| **Male; N (%)** | 8538 (38.8%) | 85,738 (41.1%) |
| **Race / Ethnicity; N (%)** |  |  |
| **White** | 20716 (94.2) | 161,739 (77.6%) |
| **Black** | 669 (3.0%) | 30,710 (14.7%) |
| **Asian** | 112 (0.5%) | 3,957 (1.9%) |
| **Hispanic or Latino** | 344 (1.6%)* | 6,759 (3.2%) |
| **Other / Unknown** | 154 (0.7%) | 5,237 (2.5%) |
| **Insurance; N (%)** |  |  |
| **Medicaid** | 358 (1.6%) | 7,549 (3.6%) |
| **Medicare** | 8582 (39.0%) | 89,312 (42.9%) |
| **Commercial** | 12,765 (58.0%) | 90,248 (43.3%) |
| **Other / Uninsured / Unknown** | 290 (1.3%) | 21,293 (10.2%) |
| **Comorbidities; N (%)** |  |  |
| **Diabetes** | 7798 (35.5%) | 59,731 (28.7%) |
| **Heart failure** | 2802 (12.7%) | 17,936 (8.6%) |
| **CAD** | 1660 (7.5%) | 35,519 (17.0%) |
| **PAD** | 47 (0.2%) | 1,721 (0.8%) |
| **MI** | 1302 (5.9%) | 6,191 (3.0%) |
| **TIA / Stroke** | 1743 (7.9%) | 9,367 (4.5%) |
| **PCI** | 563 (2.6%) | 6,012 (2.9%) |
| **CABG** | 924 (4.2%) | 3,313 (1.6%) |

\*Hispanic / Latino ethnicity recorded separately from race in Geisinger data; thus race/ethnicity categories do not sum to 100%

### Primary Outcome

The proportion of individuals with a CKD diagnosis code documented during an outpatient encounter within 1 year of the index date at Geisinger increased from 28.4% in 2008-2012 to 65.7% in 2013-2023 (p<0.001). Across years, there was a strong correlation between CKD auto-loading and CKD diagnosis (r=0.98) (**Figure 2**). Each 10 percentage-point increase in CKD auto-load was associated with 7.2 percentage-point (95% CI: 6.1%, 8.3%; p<0.001) higher absolute prevalence of CKD diagnosis documentation. By comparison, the corresponding proportion with outpatient CKD diagnosis code in the 4 OLDW health systems decreased from 22.6% in 2008-2012 to 20.7% in 2013-2023. In DiD analysis, the CKD auto-load implementation period was associated with a 39.1% (95% CI: 38.1%, 40.1%; p<0.001) absolute increase in CKD diagnosis at Geisinger compared with the 4 OLDW health systems **(Figure 3)**. Overall, CKD diagnosis occurred more frequently at Geisinger vs. OLDW and CKD diagnosis was more rapid for CKD stage 4 and 3b compared to CKD stage 3a in both cohorts (**Supplemental Figure 1**).

**Figure 3:**
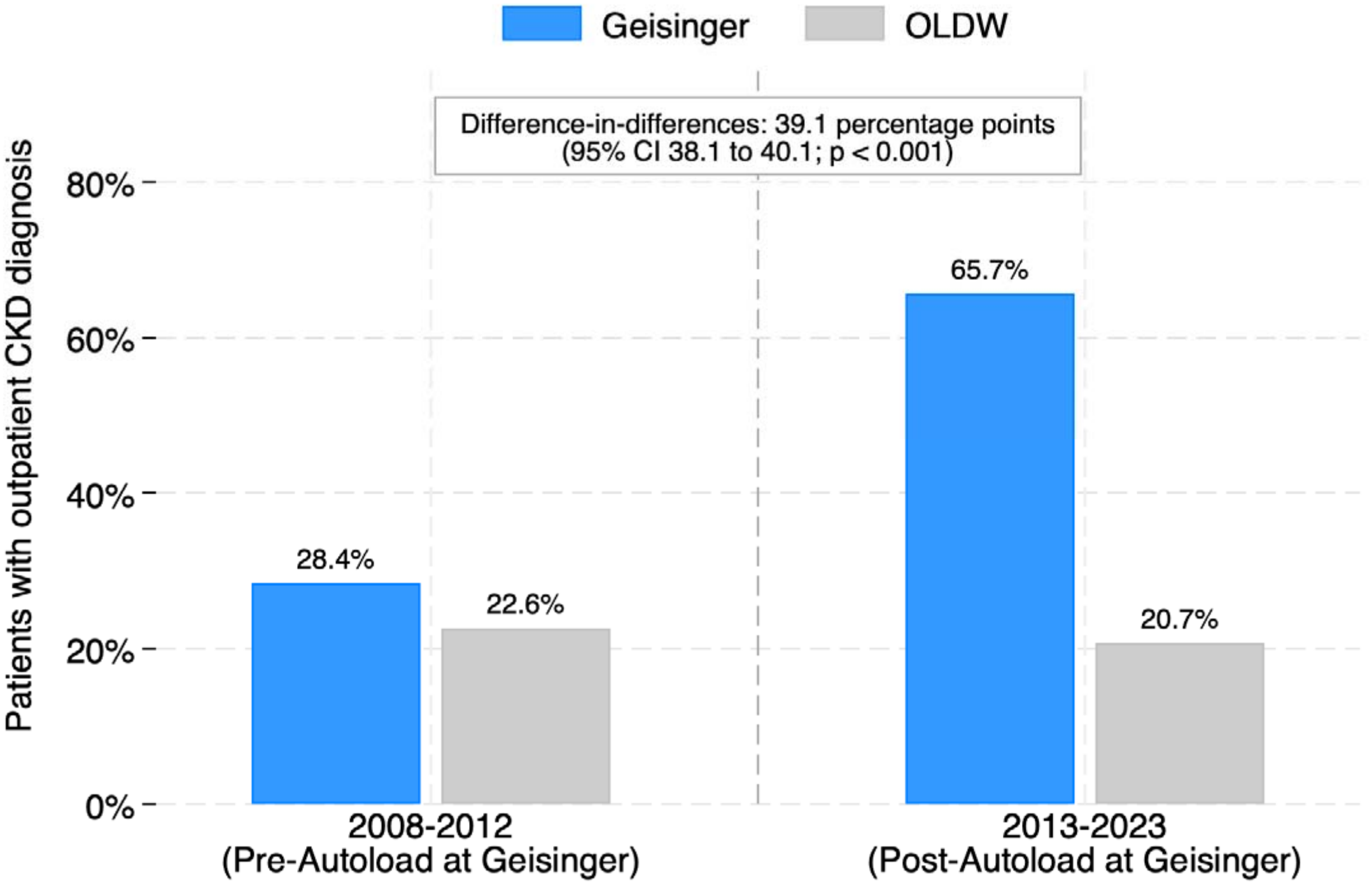
Proportion of patients with outpatient CKD diagnosis before and after implementation of CKD Autoload at Geisinger vs. OLDW

### CKD-related lab testing

Among secondary outcomes of this study, the CKD auto-load time period was associated with an increase in some but not all CKD-related labs (**Table 2, Supplemental Tables S2 and S3)**. After the CKD auto-load was instituted, measurement of follow-up eGFR (or serum creatinine) within 1 year was stable at Geisinger (pre-implementation 83.3% to post-implementation 83.6%) whereas it decreased at the 4 OLDW health systems (86.6% to 77.8%) for an absolute DiD of 9.1% (95% CI: 7.9%, 10.2%). Protein/creatinine ratio (PCR) testing increased at Geisinger relative to the 4 OLDW health systems (DiD 3.0%, 95% CI: 2.3%, 3.8%) whereas ACR testing increased more at OLDW than at Geisinger (-3.0%, 95% CI: -4.3%, -1.6%).

**Table 2.** Difference-in-Differences (DiD) Analyses for Proportions of Individuals with CKD Lab Testing within 1 Year of Index Date of Lab-based Incident CKD.

| Outcome | Geisinger Pre/Post | Δ (SE) | OLDW Pre/Post | Δ (SE) | DiD (95% CI) | p value |
|---|---|---|---|---|---|---|
| eGFR (or serum creatinine) | 83.3 / 83.6 | 0.3 (0.6) | 86.6 / 77.8 | −8.8 (0.2) | 9.1 (7.9, 10.2) | <0.001 |
| Albumin/creatinine ratio | 29.9 / 28.9 | −1.0 (0.7) | 19.4 / 21.4 | 2.0 (0.2) | −3.0 (−4.3, −1.6) | <0.001 |
| Protein/creatinine ratio | 5.8 / 9.2 | 3.4 (0.4) | 3.5 / 3.9 | 0.4 (0.1) | 3.0 (2.3, 3.8) | <0.001 |
| Phosphorus | 20.3 / 42.5 | 22.2 (0.6) | 15.2 / 12.3 | −2.9 (0.2) | 25.1 (23.8, 26.4) | <0.001 |
| Parathyroid hormone | 8.9 / 19.4 | 10.5 (0.5) | 8.0 / 6.5 | −1.5 (0.1) | 11.9 (11.0, 12.9) | <0.001 |
| 25-Hydroxyvitamin D | 27.6 / 20.3 | −7.3 (0.6) | 18.8 / 27.6 | 8.8 (0.2) | −16.2 (−17.5, −14.8) | <0.001 |
| Hemoglobin | 67.4 / 69.2 | 1.8 (0.7) | 82.4 / 84.4 | 2.1 (0.2) | −0.3 (−1.7, 1.1) | 0.67 |

The association of CKD auto-loading with CKD-related lab testing was examined using a difference-in-differences (DiD) design, comparing lab testing 12 months after index date of incident CKD post-implementation vs. pre-implementation.

The most prominent DiD estimates associated with the CKD auto-load were for CKD bone mineral disease labs. Phosphorus testing more than doubled at Geisinger (20.3% to 42.5%) vs. slight decrease at OLDW health systems (15.2% to 12.3%) for an absolute DiD of 25.1% (95% CI: 23.8%, 26.4%), and parathyroid hormone testing also more than doubled at Geisinger (8.9% to 19.4%) vs. slight decrease at OLDW health systems (8.0% to 6.5%) for an absolute DiD of 11.9% (95% CI: 11.0%, 12.9%). However, lab testing for 25-hydroxyvitamin D decreased at Geisinger from 27.6% to 20.3% whereas it increased at the 4 OLDW health systems from 18.8% to 27.6% for an absolute DiD of -16.2% (95% CI: -17.5%, -14.8%).

#### Guideline-directed medical therapy

Overall, baseline use of ACEi/ARB and statins was substantially higher at Geisinger compared to the 4 OLDW health systems throughout all time periods (**Figure 4, Tables S4-S6)**. At Geisinger, ACEi/ARB use increased slightly from 64% to 65% (12 months before vs. 12 months after lab-based incident CKD) in the pre-intervention period and from 61% to 64% in the post-intervention period, compared with increases from 29% to 37% in the pre-intervention period and from 44% to 45% in the post-intervention period in the OLDW health systems. Similarly, statin use among those >50 years of age increased from 63% to 66% in the pre-intervention period and from 62% to 67% in the post-intervention period at Geisinger, compared with increases from 25% to 34% and from 40% to 43%, respectively, in the OLDW health systems. SGLT2i use increased over time, with use at Geisinger increasing from 2% to 3% in the pre-guideline period (2014-2020) and from 6% to 11% in the post-guideline period (2021-2023), compared with increases from 1% to 2% in the pre-guideline period and from 5% to 7% in the post-guideline period in the OLDW health systems.

**Figure 4:**
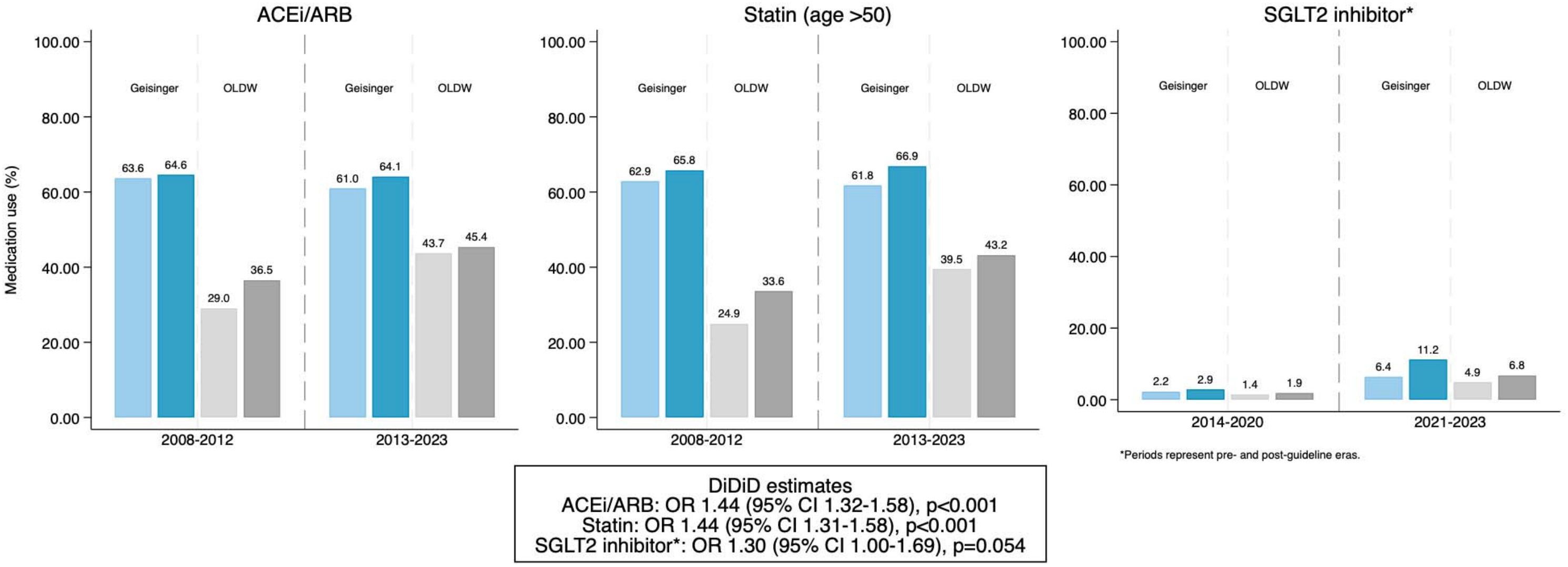
Difference-in-difference-in-differences Analyses for Medication use. Difference-in-difference-in-differences (DiDiD) analyses compared GDMT use during the 12 months after vs. 12 months before incident lab-defined CKD between Geisinger and the four OLDW health systems. For ACEi/ARB and statin use (among adults ≥50 years), changes were compared between the pre-implementation (2008-2012) and routine CKD auto-load (2013-2023) periods. For SGLT2 inhibitor use, an exploratory guideline-era analysis compared 2021-2023 with 2014-2020 since SGLT2 inhibitors were not available during the pre-implementation period. The DiDiD odds ratios and 95% confidence intervals represent the three-way interaction assessing whether the pre- to-post incident lab-based CKD change in medication use differed between the pre- and post-time periods at Geisinger beyond temporal changes observed at OLDW.

In DiDiD analyses, the CKD auto-load intervention time period was associated with higher post-incident CKD uptake of ACEi/ARB, SGLT2i and statins among those age >50 years at Geisinger, compared to OLDW health systems. The CKD auto-load was associated with OR 1.44 (95% CI 1.32 -1.58; p<0.001) for ACEi/ARB use, OR 1.44 (95% CI: 1.31-1.58; p<0.0001) for statins for those >50 years, and OR 1.30 (95% CI 1.00-1.69; p=0.05) for SGLT2 inhibitors (**Figure 4, Tables S4-S6**).

GLP1 RA use and nsMRA use are shown in **Table S5**, but DiDiDs were not calculated due to lack of guidelines specific to CKD for GLP-1 RA during the time period and very low use of nsMRA in all groups (<1%).

## Discussion

This retrospective analysis evaluates the impact of an algorithm-based automatic loading of CKD diagnosis in the EHR problem list of patients who met criteria for the laboratory-based diagnosis of CKD and who were receiving primary or nephrology outpatient care at a large health system. When compared with four similarly sized OLDW health systems, we found that among patients with incident and undiagnosed CKD, CKD diagnosis in outpatient encounters within 1 year increased from 28% in 2008-2012 to 66% in 2013-2023 at Geisinger while it decreased from 23% to 21% at the comparison health systems. CKD auto-load was associated with increases in completion of some CKD-related labs and increased use of CKD GDMT.

Our findings extend upon prior studies showing suboptimal documentation of CKD diagnosis in the EHR. For instance, one large health system reported that 11% had CKD documented in the problem list between 2005-2009.^5^ Another study found that 16% of patients with stage 3 or 4 CKD had documentation of CKD on the problem list between 2007-2008.^6^ Finally, in a study of 2010-2011 Medicare patients, only 12% who met laboratory criteria for CKD had clinically recognized CKD diagnosis.^18^ One longitudinal analysis between 2009 to 2020 found that the median time from laboratory-based diagnosis of CKD until the first ICD code for CKD in the EHR-linked claims data was 3.6 years (interquartile range: 1.0-8.4 years).^7^ Our study showed that the CKD auto-load process was significantly associated with improvements in diagnosis. Prior literature shows that higher CKD documentation is associated with improved CKD-specific testing^5^.

In recent years, several initiatives have been undertaken in a variety of settings to improve earlier diagnosis of CKD.^19^ Emphasis, in particular, has been placed on clinical decision support tools in the EHR to improve CKD diagnosis.^20-22^ Use of EHR-based algorithms to diagnose CKD has been found to be effective.^23,24^ Our study is, to our knowledge, the first to report the effectiveness of automatic loading of CKD diagnosis to EHR problem list of patients who meet lab criteria for CKD diagnosis, resulting in improved disease burden capture. Accurate patient problem lists can enable better continuity of care, facilitate closer surveillance, and are expected to be associated with greater guideline-directed interventions. We also find that improved CKD diagnosis is associated with some improvements in CKD-related lab ordering and GDMT use that could potentially lead to improved future kidney and cardiovascular outcomes. Disparate findings for some of the CKD-related labs demonstrate that there are other factors that can impact testing. The lower rates of vitamin D screening occurring at Geisinger in later years may be attributed to a Geisinger-wide Choosing Wisely initiate (an American Board of Internal Medicine Foundation campaign) that encouraged providers to decrease vitamin D testing in the overall population.

In addition to clinical care, complete problem list documentation is also important for the Centers for Medicare and Medicaid Services (CMS) risk adjustment. CKD is a high-impact chronic condition included in multiple risk adjustment methodologies, including the CMS Hierarchical Condition Category (HCC) model and Medicare Advantage risk adjustment. Risk scores are calculated from documented diagnoses rather than laboratory values alone. Consequently, patients with laboratory-confirmed CKD who lack a documented diagnosis may have their disease burden underestimated, leading to lower risk-adjustment reimbursement despite requiring more complex care. However, our study shows that automatic loading of CKD diagnosis is associated with relatively modest increases in GDMT use. This highlights that while improved documentation and recognition is important, other actions are needed to influence downstream clinical outcomes. Automatic loading of diagnoses will likely need to be combined with other interventions like CDS alerts, population health management, and team-based integrated care models. Furthermore, to preserve trust and minimize inappropriate labeling, auto-loading algorithms would need to continue improving diagnostic specificity through incorporation of accepted diagnostic criteria with clinical oversight and possibly provider confirmation.

The strengths of this study include a large population of CKD patients in the intervention health system achieving a high rate of disease burden capture, and use of comparator health systems to account for temporal changes. There were several limitations in our study. First, we were only able to do summary-level comparisons between Geisinger and OLDW, and thus we could not make adjustments for differences in characteristics between sites. Regardless, the DiD design attempts to mitigate time-invariant between-system differences. The implementation of the CKD auto-load did not occur uniformly at one specific moment, but rather was implemented and used less frequently from 2008-2012, and then much more frequently in 2013 and onwards. Thus, estimates of the effect of the CKD auto-load implementation are likely underestimates of what could happen if CKD auto-loading were implemented fully in a health system, compared with no baseline use at all. There may also have been other changes ongoing at the health system level simultaneous to autoload (e.g. changes to CDS tools, ongoing PCP education) that make it harder to ascribe any causality. We also have no knowledge of any changes in CDS or system-level educational efforts around CKD that may have occurred at the comparator OLDW health systems. Finally, we look at medication prescriptions and do not evaluate actual fills.

## Conclusions

An algorithm-based problem list auto-load process for CKD was associated with increased CKD diagnosis, completion of some CKD-related labs, and improved use of GDMT. This suggests that CKD auto-loading is a practical, scalable strategy to improve clinical recognition of CKD and supports evidence-based monitoring, enhances the accuracy of CMS risk adjustment, and strengthens health system performance within value-based reimbursement models.

## Data Availability

Some of the data in the study belong to a third party (Optum Labs Data Warehouse), and authors are not authorized to share the data.

