## Supplemental Tables for "Improvement in Chronic Kidney Disease (CKD) diagnosis and management after automatic loading of CKD in the electronic health record problem list"

**Table S1**: International Classification of Diseases Ninth or Tenth revision (ICD 9/10) codes used in the study

| **Comorbid conditions** | **Codes** |
| --- | --- |
| CKD | 585.3 CKD stage 3, 585.4 CKD stage 4, N18.30 CKD stage 3 unspecified, N18.31 CKD stage 3a, N18.32 CKD stage 3b, N18.4 CKD stage 4, E11.21 T2DM with diabetic nephropathy; E11.22 T2DM with diabetic CKD; I12.9 Hypertensive CKD with stages 1-4 CKD or unspecified CKD |
| Heart failure | ICD 9: 428  ICD 10: I42.5-I42.9, I43, I50, I09.9, I11.0, I13.0, I13.2, I25.5, I42.0, P29.0 |
| Coronary artery disease | ICD 9: 412, 414.0, 414.1, 414.2, 414.3, 414.8, 414.9, 429.2, 793.2  ICD 10: I25.1, I25.2, I25.5, I25.6, I25.70-I25.76, I25.79, I25.8, I25.9, R93.1 |
| Stroke/transient ischemic attack | ICD 9: 430, 431, 434.01, 434.11, 434.91, 436, 433, 415.1, 444, 435  ICD 10: G46.0-2, I26.01, I26.02, I26.09, I26.92, I26.99, I67.841, I67.848, I67.89, G45, I60, I61, I63, I74 |
| Peripheral artery disease | ICD 9: 440.2, 440.20, 440.21, 440.22, 440.23, 440.24, 440.29, 440.3, 440.4, 38.18, 39.25, 39.29, 39.50  ICD 10: I70.2, I70.20, I70.21, I70.22, I70.23, I70.24, I70.25, I70.26, I70.29, I70.3, I70.4, I70.5, I70.92, 0312096-031209C, 03120A6-03120AC, 03120J6-03120JC, 03120K6-03120KC, 03120Z6-03120ZC, 0313096-031309C, 03130A6-03130AC, 03130J6-03130JC, 03130K6-03130KC, 03130Z6-03130ZC, 0314096-031409C, 03140A6-03140AC, 03140J6-03140JC, 03140K6-03140KC, 03140Z6-03140ZC, 0315096-031509C, 03150A6-03150AC, 03150J6-03150JC, 03150K6-03150KC, 03150Z6-03150ZC, 0316096-031609C, 03160A6-03160AC, 03160J6-03160JC, 03160K6-03160KC, 03160Z6-03160ZC, 041009H-041009R, 04100AH-04100AR, 04100JH-04100JR, 04100KH-04100KR, 04100ZH-04100ZR, 041C09H-041C09R, 041C0AH-041C0AR, 041C0JH-041C0JR, 041C0KH-041C0KR, 041C0ZH-041C0ZR, 041D09H-041D09R, 041D0AH-041D0AR, 041D0JH-041D0JR, 041D0KH-041D0KR, 041D0ZH-041D0ZR, 041E09H-041E09Q, 041E0AH-041E0AQ, 041E0JH-041E0JQ, 041E0KH-041E0KQ, 041E0ZH-041E0ZQ, 041F09H-041F09Q, 041F0AH-041F0AQ, 041F0JH-041F0JQ, 041F0KH-041F0KQ, 041F0ZH-041F0ZQ, 041H09H-041H09Q, 041H0AH-041H0AQ, 041H0JH-041H0JQ, 041H0KH-041H0KQ, 041H0ZH-041H0ZQ, 041J09H-041J09Q, 041J0AH-041J0AQ, 041J0JH-041J0JQ, 041J0KH-041J0KQ, 041J0ZH-041J0ZQ, 041K, 041L, 041M, 041N, 041P, 041Q, 041R, 041S, 041T, 041U, 041V, 041W, 047C, 047D, 047E, 047F, 047H, 047J, 047K, 047L, 047M, 047N, 047P, 047Q, 047R, 047S, 047T, 047U, 047V, 047W, 047Y, 04BC, 04BD, 04BE, 04BF, 04BH, 04BJ, 04BK, 04BL, 04BM, 04BN, 04BP, 04BQ, 04BR, 04BS, 04BT, 04BU, 04BV, 04BW, 04BY, 04CC, 04CD, 04CE, 04CF, 04CH, 04CJ, 04CK, 04CL, 04CM, 04CN, 04CP, 04CQ, 04CR, 04CS, 04CT, 04CU, 04CV, 04CW, 04CY, 04HC0DZ, 04HC3DZ, 04HD0DZ, 04HD3DZ,04HE0DZ, 04HE3DZ,04HF0DZ, 04HF3DZ,04HH0DZ, 04HH3DZ,04HJ0DZ, 04HJ3DZ,04HK0DZ, 04HK3DZ,04HL0DZ, 04HL3DZ,04HM0DZ, 04HM3DZ,04HN0DZ, 04HN3DZ,04HP0DZ, 04HP3DZ,04HQ0DZ, 04HQ3DZ,04HR0DZ, 04HR3DZ,04HS0DZ,04HS3DZ,04HT0DZ, 04HT3DZ,04HU0DZ, 04HU3DZ,04HV0DZ, 04HV3DZ,04HW0DZ, 04HW3DZ,04HY0DZ, 04HY3DZ, 04RC, 04RD, 04RE, 04RF, 04RH, 04RJ, 04RK, 04RL,04RM, 04RN, 04RP, 04RQ, 04RR, 04RS, 04RT, 04RU, 04RV, 04RW, 04RY, 04UC, 04UD, 04UE, 04UF, 04UH, 04UJ,04UK, 04UL, 04UM, 04UN, 04UP, 04UQ, 04UR, 04US, 04UT, 04CU, 04UV, 04UW, 04UY, 0Y06 |
| Myocardial infarction | ICD 9: 410, 412  ICD 10: I21, I22, I25.2 |
| Unstable angina | ICD 9: 411.1  ICD 10: I20.0 |
| Percutaneous coronary intervention | ICD 9: V45.82, 00.66, 17.55, 36.01, 36.02, 36.05, 36.06, 36.07, 36.09  ICD 10: Z95.5, Z98.61, 0270046, 027004Z, 02700D6, 02700DZ, 02700T6, 02700TZ, 02700Z6, 02700ZZ, 0270346, 027034Z, 02703D6, 02703DZ, 02703T6, 02703TZ, 02703Z6, 02703ZZ, 0270446, 027044Z, 02704D6, 02704DZ, 02704T6, 02704TZ, 02704Z6, 02704ZZ, 0271046, 027104Z, 02710D6, 02710DZ, 02710T6, 02710TZ, 02710Z6, 02710ZZ, 0271346, 027134Z, 02713D6, 02713DZ, 02713T6, 02713TZ, 02713Z6, 02713ZZ, 0271446, 027144Z, 02714D6, 02714DZ, 02714T6, 02714TZ, 02714Z6, 02714ZZ, 0272046, 027204Z, 02720D6, 02720DZ, 02720T6, 02720TZ, 02720Z6, 02720ZZ, 0272346, 027234Z, 02723D6, 02723DZ, 02723T6, 02723TZ, 02723Z6, 02723ZZ, 0272446, 027244Z, 02724D6, 02724DZ, 02724T6, 02724TZ, 02724Z6, 02724ZZ, 0273046, 027304Z, 02730D6, 02730DZ, 02730T6, 02730TZ, 02730Z6, 02730ZZ, 0273346, 027334Z, 02733D6, 02733DZ, 02733T6, 02733TZ, 02733Z6, 02733ZZ, 0273446, 027344Z, 02734D6, 02734DZ, 02734T6, 02734TZ, 02734Z6, 02734ZZ, 02C03ZZ, 02C04ZZ, 02C13ZZ, 02C14ZZ, 02C23ZZ, 02C24ZZ, 02C33ZZ, 02C34ZZ, X2C0361, X2C1361  HCPCS/CPT: 92920, 92921, 92924, 92925, 92928, 92929, 92933, 92934, 92937, 92938, 92941, 92943, 92944, 92980, 92981, 92982, 92984, 92995, 92996, C9600, C9601, C9602, C9603, C9604, C9605, C9606, C9607, C9608, G0290, G0291 |
| Coronary artery bypass grafting | ICD 9: V45.81, 36.10, 36.11, 36.12, 36.13, 36.14, 36.15, 36.16, 36.17, 36.19  ICD 10: Z95.1, 0210093, 0210098, 0210099, 021009C, 021009F, 021009W, 02100A3, 02100A8, 02100A9, 02100AC, 02100AF, 02100AW, 02100J3, 02100J8, 02100J9, 02100JC, 02100JF, 02100JW, 02100K3, 02100K8, 02100K9, 02100KC, 02100KF, 02100KW, 02100Z3, 02100Z8, 02100Z9, 02100ZC, 02100ZF, 0210344, 02103D4, 0210444, 0210493, 0210498, 0210499, 021049C, 021049F, 021049W, 02104A3, 02104A8, 02104A9, 02104AC, 02104AF, 02104AW, 02104D4, 02104J3, 02104J8, 02104J9, 02104JC, 02104JF, 02104JW, 02104K3, 02104K8, 02104K9, 02104KC, 02104KF, 02104KW, 02104Z3, 02104Z8, 02104Z9, 02104ZC, 02104ZF, 0211093, 0211098, 0211099, 021109C, 021109F, 021109W, 02110A3, 02110A8, 02110A9, 02110AC, 02110AF, 02110AW, 02110J3, 02110J8, 02110J9, 02110JC, 02110JF, 02110JW, 02110K3, 02110K8, 02110K9, 02110KC, 02110KF, 02110KW, 02110Z3, 02110Z8, 02110Z9, 02110ZC, 02110ZF, 0211344, 02113D4, 0211444, 0211493, 0211498, 0211499, 021149C, 021149F, 021149W, 02114A3, 02114A8, 02114A9, 02114AC, 02114AF, 02114AW, 02114D4, 02114J3, 02114J8, 02114J9, 02114JC, 02114JF, 02114JW, 02114K3, 02114K8, 02114K9, 02114KC, 02114KF, 02114KW, 02114Z3, 02114Z8, 02114Z9, 02114ZC, 02114ZF, 0212093, 0212098, 0212099, 021209C, 021209F, 021209W, 02120A3, 02120A8, 02120A9, 02120AC, 02120AF, 02120AW, 02120J3, 02120J8, 02120J9, 02120JC, 02120JF, 02120JW, 02120K3, 02120K8, 02120K9, 02120KC, 02120KF, 02120KW, 02120Z3, 02120Z8, 02120Z9, 02120ZC, 02120ZF, 0212344, 02123D4, 0212444, 0212493, 0212498, 0212499, 021249C, 021249F, 021249W, 02124A3,02124A8, 02124A9, 02124AC, 02124AF, 02124AW, 02124D4, 02124J3, 02124J8, 02124J9, 02124JC, 02124JF, 02124JW, 02124K3, 02124K8, 02124K9, 02124KC, 02124KF, 02124KW, 02124Z3, 02124Z8, 02124Z9, 02124ZC, 02124ZF, 0213093, 0213098, 0213099, 021309C, 021309F, 021309W, 02130A3, 02130A8, 02130A9, 02130AC, 02130AF, 02130AW, 02130J3, 02130J8, 02130J9, 02130JC, 02130JF, 02130JW, 02130K3, 02130K8, 02130K9, 02130KC, 02130KF, 02130KW, 02130Z3, 02130Z8, 02130Z9, 02130ZC, 02130ZF, 0213344, 02133D4, 0213444, 0213493, 0213498, 0213499, 021349C, 021349F, 021349W, 02134A3, 02134A8, 02134A9, 02134AC, 02134AF, 02134AW, 02134D4, 02134J3, 02134J8, 02134J9, 02134JC, 02134JF, 02134JW, 02134K3, 02134K8, 02134K9, 02134KC, 02134KF, 02134KW, 02134Z3, 02134Z8, 02134Z9, 02134ZC, 02134ZF  HCPCS/CPT: 33510, 33511, 33512, 33513, 33514, 33516, 33517, 33518, 33519, 33521, 33522, 33523, 33533, 33534, 33535, 33536 |

**Table S2** shows the proportions of completed CKD-related labs within 12 months of the index date of lab-based CKD at Geisinger compared to health systems within the OLDW data.

| Year | **Number of patients** | | **Creatinine** | | **Estimated GFR** | | **Albumin to Creatinine ratio (ACR)** | | **Protein to Creatinine ratio (PCR)** | |
| --- | --- | --- | --- | --- | --- | --- | --- | --- | --- | --- |
|  | Geisinger | OLDW | Geisinger | OLDW | Geisinger | OLDW | Geisinger | OLDW | Geisinger | OLDW |
| 2008 | 1047 | 3,210 | 89% | 91% | 89% | 91% | 36% | 16% | 5% | 4% |
| 2009 | 1365 | 6,977 | 83% | 84% | 83% | 84% | 32% | 13% | 6% | 2% |
| 2010 | 1226 | 4,495 | 83% | 83% | 83% | 83% | 31% | 17% | 6% | 2% |
| 2011 | 1510 | 8,283 | 83% | 89% | 83% | 89% | 28% | 23% | 6% | 5% |
| 2012 | 1355 | 16,163 | 80% | 87% | 80% | 87% | 25% | 22% | 6% | 4% |
| 2013 | 1391 | 20,735 | 82% | 84% | 82% | 84% | 27% | 22% | 7% | 4% |
| 2014 | 1303 | 19,683 | 84% | 81% | 84% | 81% | 30% | 22% | 6% | 3% |
| 2015 | 1346 | 21,736 | 86% | 81% | 87% | 81% | 31% | 21% | 9% | 3% |
| 2016 | 1813 | 19,049 | 85% | 80% | 85% | 80% | 27% | 21% | 10% | 3% |
| 2017 | 1622 | 18,084 | 82% | 77% | 83% | 77% | 24% | 22% | 11% | 4% |
| 2018 | 1554 | 16,392 | 84% | 78% | 85% | 78% | 27% | 22% | 12% | 4% |
| 2019 | 1670 | 16,079 | 83% | 66% | 84% | 66% | 25% | 20% | 11% | 4% |
| 2020 | 1608 | 10,638 | 81% | 76% | 82% | 76% | 27% | 21% | 11% | 5% |
| 2021 | 1428 | 10,636 | 80% | 75% | 80% | 75% | 30% | 21% | 8% | 5% |
| 2022 | 956 | 8,852 | 81% | 74% | 81% | 74% | 39% | 22% | 7% | 5% |
| 2023 | 801 | 7,390 | 83% | 77% | 83% | 77% | 43% | 24% | 9% | 6% |

**Table S3** shows the proportions of completed CKD-related labs within 12 months of the index date of lab-based CKD at Geisinger compared to health systems within the OLDW data.

|  | **Intact parathyroid hormone** | | **Phosphorus** | | **25-hydroxyvitamin D** | | **Hemoglobin** | | **Cholesterol panel** | |
| --- | --- | --- | --- | --- | --- | --- | --- | --- | --- | --- |
|  | **Geisinger** | **OLDW** | **Geisinger** | **OLDW** | **Geisinger** | **OLDW** | **Geisinger** | **OLDW** | **Geisinger** | **OLDW** |
| 2008 | 10% | 8% | 17% | 18% | 0% | 11% | 74% | 82% | 78% | 69% |
| 2009 | 9% | 6% | 19% | 15% | 0% | 11% | 73% | 81% | 77% | 66% |
| 2010 | 9% | 5% | 27% | 9% | 0% | 15% | 71% | 79% | 77% | 70% |
| 2011 | 9% | 11% | 18% | 17% | 0% | 21% | 61% | 84% | 60% | 76% |
| 2012 | 8% | 8% | 20% | 15% | 0% | 24% | 60% | 83% | 56% | 76% |
| 2013 | 13% | 7% | 26% | 14% | 0% | 24% | 64% | 82% | 55% | 77% |
| 2014 | 20% | 6% | 31% | 12% | 0% | 23% | 65% | 82% | 57% | 78% |
| 2015 | 34% | 6% | 46% | 12% | 0% | 24% | 70% | 83% | 60% | 76% |
| 2016 | 29% | 6% | 42% | 12% | 0% | 26% | 69% | 84% | 59% | 74% |
| 2017 | 20% | 6% | 36% | 13% | 0% | 28% | 68% | 85% | 55% | 74% |
| 2018 | 20% | 6% | 47% | 13% | 0% | 27% | 73% | 86% | 57% | 75% |
| 2019 | 13% | 6% | 50% | 10% | 0% | 26% | 71% | 83% | 54% | 75% |
| 2020 | 10% | 7% | 49% | 12% | 11% | 33% | 70% | 89% | 55% | 79% |
| 2021 | 1% | 7% | 46% | 12% | 18% | 35% | 66% | 88% | 56% | 77% |
| 2022 | 0% | 7% | 46% | 12% | 19% | 36% | 71% | 89% | 54% | 76% |
| 2023 | 0% | 8% | 51% | 14% | 25% | 38% | 76% | 90% | 58% | 79% |

**Table S4** Proportions of individuals with prescriptions for ACEi / ARB and SGLT2i in the 12 months before and after index date of lab-based CKD at Geisinger and within OLDW health systems

| **Year** | **ACEi or ARB one year before index date** | | **ACEi or ARB one year after index date** | | **SGLT2i one year before index date** | | **SGLT2i one year after index date** | |
| --- | --- | --- | --- | --- | --- | --- | --- | --- |
|  | Geisinger | OLDW | Geisinger | OLDW | Geisinger | OLDW | Geisinger | OLDW |
| 2008 | 69% | 23% | 68% | 29% | 0% | 0% | 0% | 0% |
| 2009 | 65% | 28% | 66% | 30% | 0% | 0% | 0% | 0% |
| 2010 | 64% | 34% | 65% | 34% | 0% | 0% | 0% | 0% |
| 2011 | 62% | 26% | 64% | 37% | 0% | 0% | 0% | 0% |
| 2012 | 60% | 31% | 61% | 41% | 0% | 0% | 0% | 0% |
| 2013 | 62% | 42% | 65% | 49% | 0% | 0% | 0% | 0% |
| 2014 | 62% | 45% | 63% | 48% | 0% | 0% | 0% | 1% |
| 2015 | 65% | 43% | 66% | 44% | 1% | 1% | 1% | 1% |
| 2016 | 65% | 43% | 67% | 43% | 1% | 1% | 2% | 2% |
| 2017 | 61% | 39% | 64% | 40% | 2% | 2% | 2% | 2% |
| 2018 | 60% | 41% | 63% | 48% | 3% | 2% | 4% | 2% |
| 2019 | 60% | 48% | 65% | 45% | 4% | 2% | 5% | 3% |
| 2020 | 57% | 47% | 60% | 47% | 4% | 3% | 5% | 4% |
| 2021 | 61% | 47% | 66% | 47% | 5% | 4% | 8% | 5% |
| 2022 | 57% | 46% | 61% | 45% | 7% | 5% | 12% | 7% |
| 2023 | 61% | 43% | 65% | 43% | 9% | 6% | 16% | 9% |

**Table S5**. Proportions of individuals with prescriptions for GLP1 agonists and statins in the 12 months before and after index date of lab-based CKD at Geisinger and within OLDW health systems

| **Year** | **GLP-1 agonists one year before index date** | | **GLP1 agonists one year after index date** | | **Statin one year before index date** | | **Statin one year after index date** | | **nsMRA one year before index date** | | **nsMRA one year after index date** | |
| --- | --- | --- | --- | --- | --- | --- | --- | --- | --- | --- | --- | --- |
|  | Geisinger | OLDW | Geisinger | OLDW | Geisinger | OLDW | Geisinger | OLDW | Geisinger | OLDW | Geisinger | OLDW |
| 2008 | 1% | 0% | 1% | 0% | 66% | 20% | 68% | 27% | 0.00% | 0.00% | 0.00% | 0.00% |
| 2009 | 0% | 0% | 1% | 0% | 61% | 23% | 64% | 26% | 0.00% | 0.00% | 0.00% | 0.00% |
| 2010 | 0% | 0% | 1% | 1% | 61% | 27% | 65% | 28% | 0.00% | 0.00% | 0.00% | 0.00% |
| 2011 | 0% | 0% | 0% | 1% | 63% | 23% | 67% | 35% | 0.00% | 0.00% | 0.00% | 0.00% |
| 2012 | 0% | 0% | 1% | 1% | 60% | 28% | 62% | 40% | 0.00% | 0.00% | 0.00% | 0.00% |
| 2013 | 1% | 0% | 1% | 1% | 62% | 39% | 65% | 46% | 0.00% | 0.00% | 0.00% | 0.00% |
| 2014 | 1% | 0% | 1% | 1% | 61% | 41% | 64% | 45% | 0.00% | 0.00% | 0.00% | 0.00% |
| 2015 | 1% | 1% | 1% | 1% | 64% | 39% | 68% | 41% | 0.00% | 0.00% | 0.00% | 0.00% |
| 2016 | 1% | 1% | 2% | 2% | 62% | 38% | 66% | 40% | 0.00% | 0.00% | 0.00% | 0.00% |
| 2017 | 1% | 2% | 3% | 2% | 59% | 36% | 65% | 38% | 0.00% | 0.00% | 0.00% | 0.00% |
| 2018 | 2% | 2% | 4% | 3% | 59% | 37% | 65% | 46% | 0.00% | 0.00% | 0.00% | 0.00% |
| 2019 | 4% | 2% | 7% | 3% | 63% | 45% | 68% | 45% | 0.00% | 0.00% | 0.00% | 0.00% |
| 2020 | 5% | 3% | 7% | 4% | 59% | 44% | 65% | 48% | 0.00% | 0.00% | 0.00% | <0.2% |
| 2021 | 6% | 4% | 8% | 5% | 63% | 45% | 68% | 49% | 0.00% | 0.00% | 0.00% | <0.2% |
| 2022 | 9% | 5% | 11% | 7% | 58% | 46% | 67% | 48% | 0.00% | <0.2% | 0.20% | <0.2% |
| 2023 | 12% | 6% | 15% | 10% | 60% | 45% | 67% | 48% | 0.00% | <0.2% | 0.40% | <0.2% |

For OLDW data, cells <11 patients are suppressed.

**Table S6.** Difference-in-difference-in-differences (DiDiD) analyses comparing changes in GDMT usage after vs. before incident CKD diagnosis, across pre-intervention (2008-2012) and post-intervention (2013-2023) periods, between Geisinger and the 4 OLDW health systems

|  | **2008-2012**  **(1 year Pre)**  **Geisinger** | **2008-2012**  **(1 year Pre)**  **OLDW** | **2008-2012**  **(1 year Post)**  **Geisinger** | **2008-2012**  **(1 year Post)**  **OLDW** | **2013-2023**  **(1 year Pre)**  **Geisinger** | **2013-2023**  **(1 year Pre)**  **OLDW** | **2013-2023**  **(1 year Post)**  **Geisinger** | **2013-2023**  **(1 year post)**  **OLDW** | **DiD**  **(2008-2012 compared to 2013-2023 period)**  **Geisinger** | **DiD**  **(2008-2012 compared to 2013-2023 period)**  **OLDW** | **Overall**  **DiDiD**  **Odds ratio (95% CI)** | **p-value** |
| --- | --- | --- | --- | --- | --- | --- | --- | --- | --- | --- | --- | --- |
| ACEi / ARB | 63.65% | 29.02% | 64.59% | 36.53% | 60.95% | 43.70% | 64.08% | 45.37% | 2.19% | -5.84% | 1.44  (1.32, 1.58) | <0.0001 |
| Statin  (Age >50) | 62.85% | 24.92% | 65.76% | 33.62% | 61.76% | 39.50% | 66.85% | 43.18% | 2.18% | -5.03% | 1.44  (1.31, 1.58) | <0.0001 |
| SGLT2i | 2.24% | 1.44% | 2.89% | 1.85% | 6.37% | 4.88% | 11.21% | 6.77% | 4.18% | 1.48% | 1.30  (1.00, 1.69) | 0.05 |

**Supplemental Figure 1:** Kaplan-Meier estimates of CKD diagnosis-free survival following incident lab-based CKD at Geisinger and OLDW. Individuals with stage 4 CKD received a CKD diagnosis more rapidly than stage 3b and stage 3a CKD at Geisinger. In the OLDW health systems, time to stage 4 and stage 3b CKD diagnosis was more rapid than for those with stage 3a CKD.

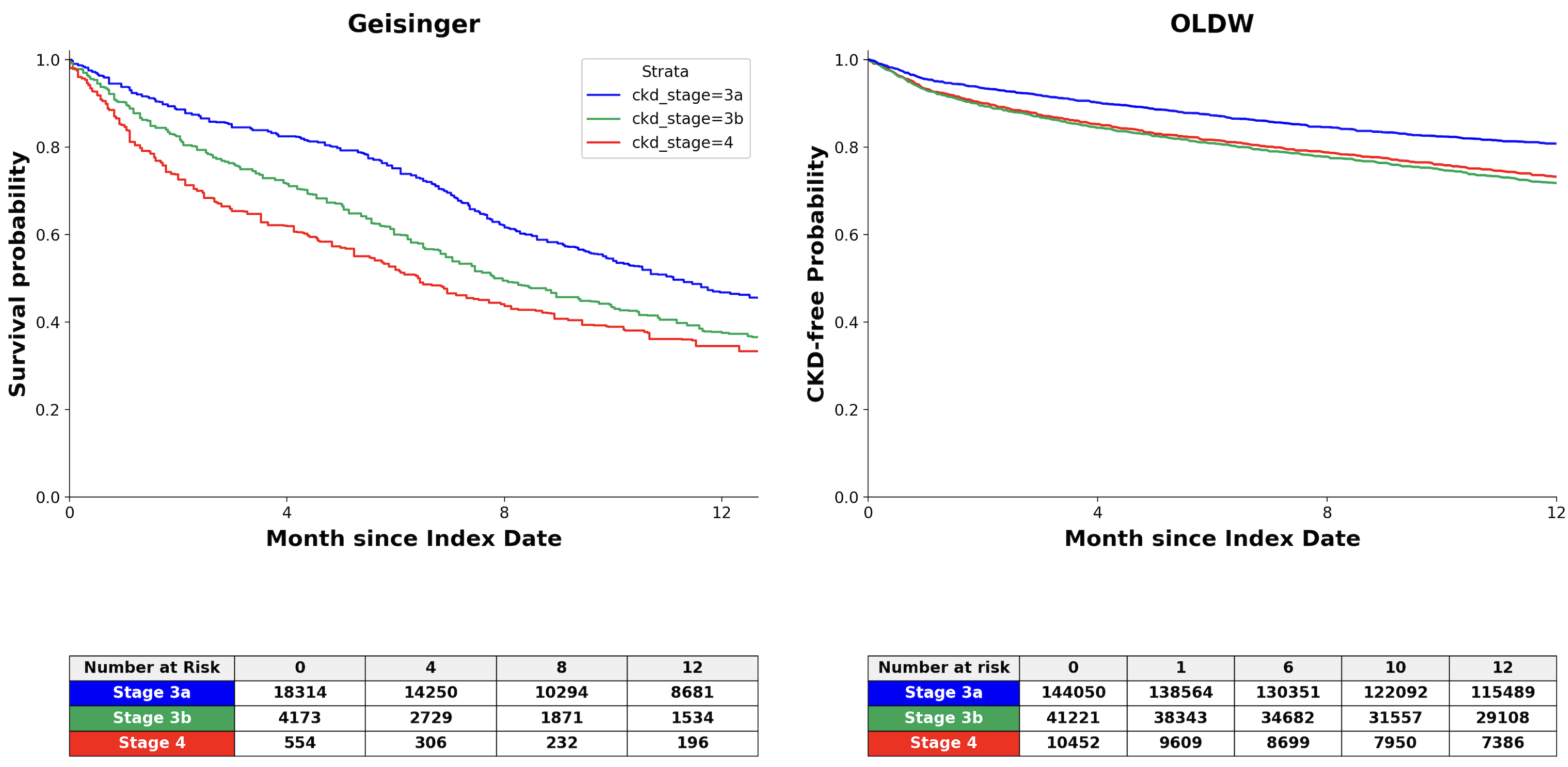
